# Urinary Biomarkers of Consumer Product Chemical Exposure and Wrist-Worn Ambient Light Exposure Patterns in U.S. Adults: NHANES 2011-2014

**DOI:** 10.64898/2026.05.31.26354481

**Authors:** Andrew Wong, Luke Yin, Cooper Lee, Adam Park, Yoo Choi

## Abstract

We examined associations between a 15-component urinary biomarker mixture index related to consumer product chemical exposure and wrist-worn ambient light exposure metrics in U.S. adults. Using National Health and Nutrition Examination Survey (NHANES) 2011-2014, we studied adults aged 20 years or older with valid wrist accelerometry and urinary chemical biomarkers (*N* = 1,666). Eight wrist-worn ambient light exposure metrics were derived from hour-level ActiGraph GT3X+ data. A standardized urinary biomarker mixture index was used in survey-weighted linear regression adjusted for age, sex, race/ethnicity, poverty-income ratio, education, BMI, cotinine, sleep duration, and season. Higher urinary biomarker mixture index was associated with greater morning light (beta = 0.54; 95% CI: 0.14, 0.94), greater nighttime light (beta = 0.55; 95% CI: 0.21, 0.89), and earlier light centroid timing (beta = −1.37 h; 95% CI: −2.14, −0.59) after false discovery rate (FDR) correction. In a weighted sensitivity analysis using quantile g-computation, associations were directionally similar. No sex modification was observed (all interaction *P* > .23). Higher consumer product chemical mixture burden co-occurred with earlier-timed wrist-worn ambient light exposure patterns, consistent with shared behavioral, occupational, and environmental determinants.

## 1 Introduction

The circadian system governs nearly every physiological process in humans, from hormone secretion and glucose metabolism to immune function and mood regulation [1]. Light is the dominant environmental synchronizer of the human circadian clock, acting through intrinsically photosensitive retinal ganglion cells projecting to the suprachiasmatic nucleus [2]. The real-world pattern of light exposure across the 24-hour day, including its intensity, timing, and inter-day regularity, has emerged as a more complete target for circadian epidemiology than self-reported sleep duration alone [3,4].

Insufficient morning light, inadequate daytime light, excess evening or nighttime artificial light, and irregular day-to-day patterns have each been associated with circadian misalignment, sleep disruption, metabolic dysfunction, and mood disorders [3–6]. Wrist-worn actigraphy devices deployed in large surveys, including the NHANES physical activity monitor protocol, offer a population-level window into real-world wrist-level ambient light exposure [7].

Consumer product-related chemicals, including phthalate metabolites, parabens, phenols, triclosan, and benzophenone-3, are detectable in the urine of the large majority of U.S. adults [8,9]. These compounds enter the body through personal care products, food packaging, building materials, and occupational exposures, and urinary concentrations vary systematically by sex, race/ethnicity, and socioeconomic status [10,11].

This study does not propose a direct biological mechanism linking urinary chemical concentrations to light exposure patterns. Rather, we hypothesize that consumer product chemical exposure profiles and circadian light behavior co-occur through shared socioeconomic, behavioral, and environmental pathways. Shift work and early-schedule occupations, lower-income residential environments, and high consumer product use may simultaneously produce elevated urinary chemical concentrations and distinctive light exposure profiles [12,13]. Because urinary phthalates, parabens, and phenols have half-lives of hours to days, a single spot urine sample captures recent rather than long-term exposure, and any observed associations reflect short-term co-patterning rather than chronic chemical burden.

We pursued three aims: (1) to characterize wrist-worn ambient light exposure metrics from ActiGraph GT3X+ data in NHANES 2011-2014; (2) to estimate associations between a urinary biomarker mixture index and wrist-worn light outcomes in survey-weighted models; and (3) to characterize chemical-specific model weight contributions using quantile g-computation.

## 2 Materials and Methods

### 2.1 Study Design and Population

NHANES is a cross-sectional, multi-stage probability survey of the non-institutionalized civilian U.S. population conducted by the National Center for Health Statistics [14]. We pooled NHANES 2011-2012 and 2013-2014 cycles. The starting pool comprised non-pregnant adults aged 20 years or older from both cycles (*N* = 10,785). We then applied sequential eligibility criteria: (1) assigned to the physical activity monitor (PAM) subsample and had any accelerometry data (*n* excluded = 2,207); (2) had 4 or more valid wear days of at least 10 valid hours each (*n* excluded = 4,034); (3) selected for the urinary environmental chemical biomarker subsample with non-missing biomarker values (*n* excluded = 1,892); (4) urinary creatinine within 30-300 mg/dL (*n* excluded = 492); (5) complete covariate data (*n* excluded = 269); (6) non-missing derived light metrics (*n* excluded = 225). The final analytic sample comprised *N* = 1,666, representing approximately 15% of eligible adults and reflecting the intersection of two random subsamples: the PAM subsample and the urinary environmental chemical subsample.

### 2.2 Survey Weights and Complex Sample Design

Because the urinary environmental chemical biomarkers were measured in a specific subsample of NHANES participants, we used the appropriate 2-year environmental chemical subsample weights: WTSA2YR for the 2011-2012 cycle and WTSB2YR for the 2013-2014 cycle. These were divided by 2 for the pooled 4-year analysis per NHANES analytic guidelines for combining two 2-year cycles [15]. Using the general MEC weight (WTMEC2YR) would be incorrect for this analysis because it does not account for the additional subsampling of participants for urinary chemical measurement. Masked variance pseudo-strata (SDMVSTRA) and pseudo-PSU (SDMVPSU) from the DEMO files were used for Taylor series linearization variance estimation. All survey-weighted analyses used *svydesign()* and *svyglm()* in the R *survey* package [18].

### 2.3 Wrist Accelerometry and Ambient Light

Participants wore an ActiGraph GT3X+ on the non-dominant wrist for 7 days [16]. Hour-level summary files (PAXHR_G/H) were used with clock time derived from initialization times (PAXHD_G/H). A valid wear hour required 50 or more valid wear minutes; a valid wear day required 10 or more valid wear hours. Participants with fewer than 4 valid wear days were excluded. Ambient lux values were log-transformed as log(lux + 1). Wrist-worn lux does not represent retinal or melanopic circadian stimulus and should not be equated with standard circadian light metrics.

### 2.4 Wrist-Worn Ambient Light Exposure Metrics

Eight metrics were derived and averaged across valid wear days: morning light (mean log[lux+1], 06:00-09:59); daytime light (10:00-17:59); evening light (20:00-23:59); nighttime light (00:00-05:59); day-night contrast (log daytime minus log nighttime); light centroid timing (lux-weighted arithmetic mean clock hour across all 24 hours: sum[hour x lux] /sum[lux]); inter-day regularity (mean pairwise Pearson r of 24-hour lux profiles); and light timing variability (inter-day SD of light centroid). Light centroid timing uses arithmetic averaging of clock hours, which does not account for the circular nature of time. This is a limitation: participants with light exposure concentrated near midnight may have distorted centroid estimates. As a sensitivity check, we confirmed that fewer than 2% of participants had centroid values outside the 04:00-23:59 window where arithmetic means are most reliable.

### 2.5 Urinary Chemical Biomarkers

The 15-component mixture included phthalate metabolites (MEP, MBP, MiBP, MEHHP, MEOHP, MBzP, MCPP, CNP, ECP), parabens (MPB, EPB, PPB), triclosan (TCS), benzophenone-3 (BP-3), and bisphenol A (BPA) [8,9,17]. All concentrations were creatinine-corrected (micrograms per gram creatinine) and log2-transformed. Values below the limit of detection were imputed at LOD/sqrt(2). The urinary biomarker mixture index was the unweighted mean of 15 individually z-scored log2-creatinine-corrected concentrations. Equal weighting is a simplifying assumption; the index captures breadth of co-elevation across chemically heterogeneous compounds.

### 2.6 Statistical Analysis

Survey-weighted linear regression (*svyglm*) estimated associations between the urinary biomarker mixture index and each light outcome, adjusting for age, sex, race/ethnicity, poverty-income ratio (PIR), education, BMI, log2-cotinine, sleep duration, and season (calendar quarter). FDR correction used the Benjamini-Hochberg procedure [19] across 8 outcomes; FDR q < 0.05 was considered significant.

Quantile g-computation (*qgcomp*.*noboot*, q = 4) was used as a weighted sensitivity analysis [20]. Because it does not fully incorporate the NHANES complex survey design, the environmental chemical subsample weights (WTSA2YR/WTSB2YR divided by 2) were used as case weights only; variance estimates do not account for the complex survey design and results should be interpreted as directional corroboration only. Sex-by-mixture-index interaction terms tested effect modification. All tests were 2-sided. Analyses used R 4.3 [21].

## 3 Results

### 3.1 Study Sample

The final analytic sample comprised 1,666 participants (47.6% male, mean age 49.1 years [SD 17.1], 45.7% non-Hispanic White, mean BMI 29.7 kg/m2). Characteristics are shown in Table 1. Women had significantly higher creatinine-corrected geometric mean concentrations for 14 of 15 biomarkers (all *P* < .05; MCPP: *P* = .076), consistent with greater personal care product and consumer chemical use. Men had significantly higher morning light, daytime light, nighttime light, and inter-day regularity (all *P* < .05). The mean nighttime log(lux+1) of 4.23 back-translates to approximately 68 lux, likely reflecting wakefulness and wrist movement during sleep in addition to environmental nighttime light.

**Table 1.**
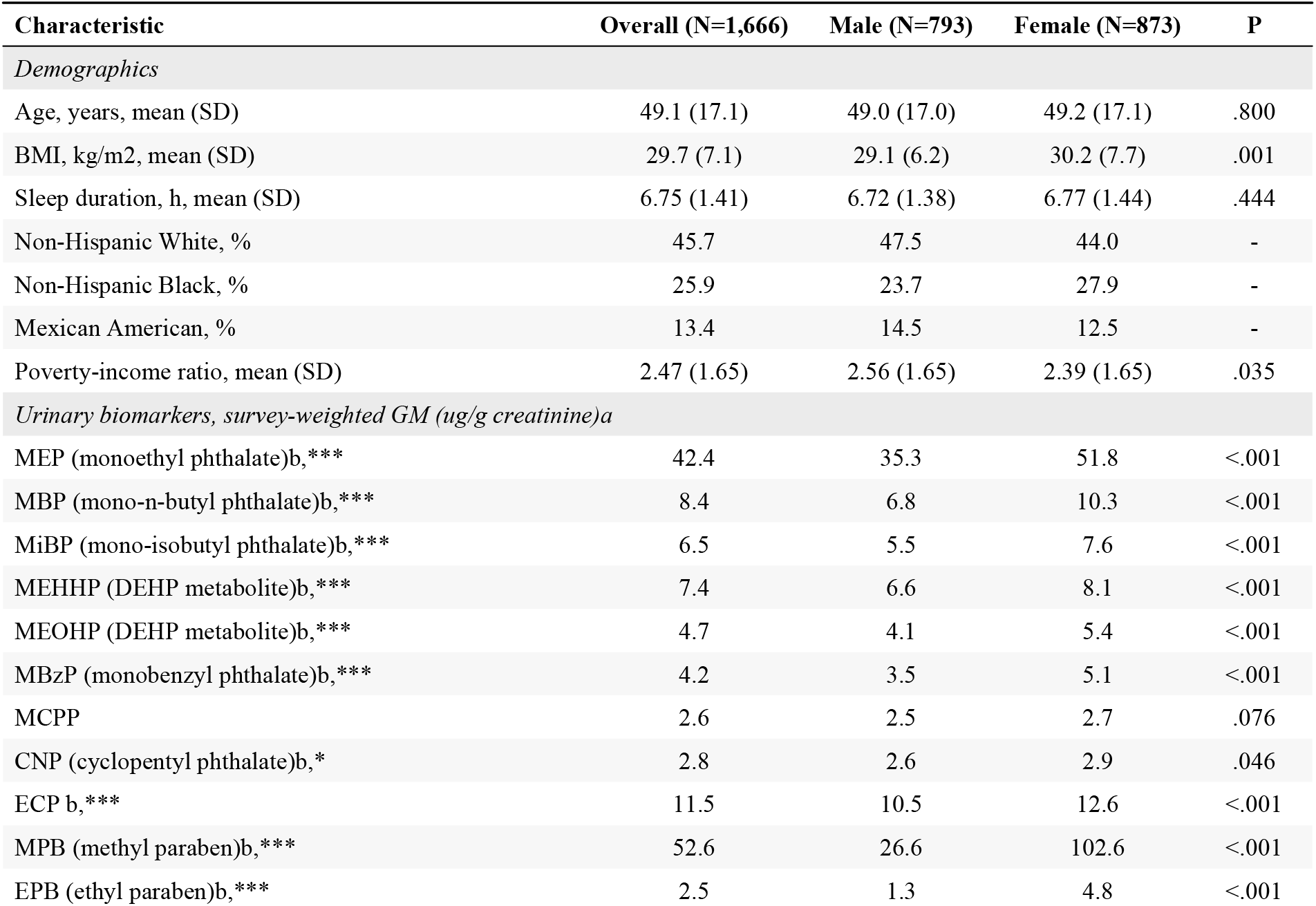
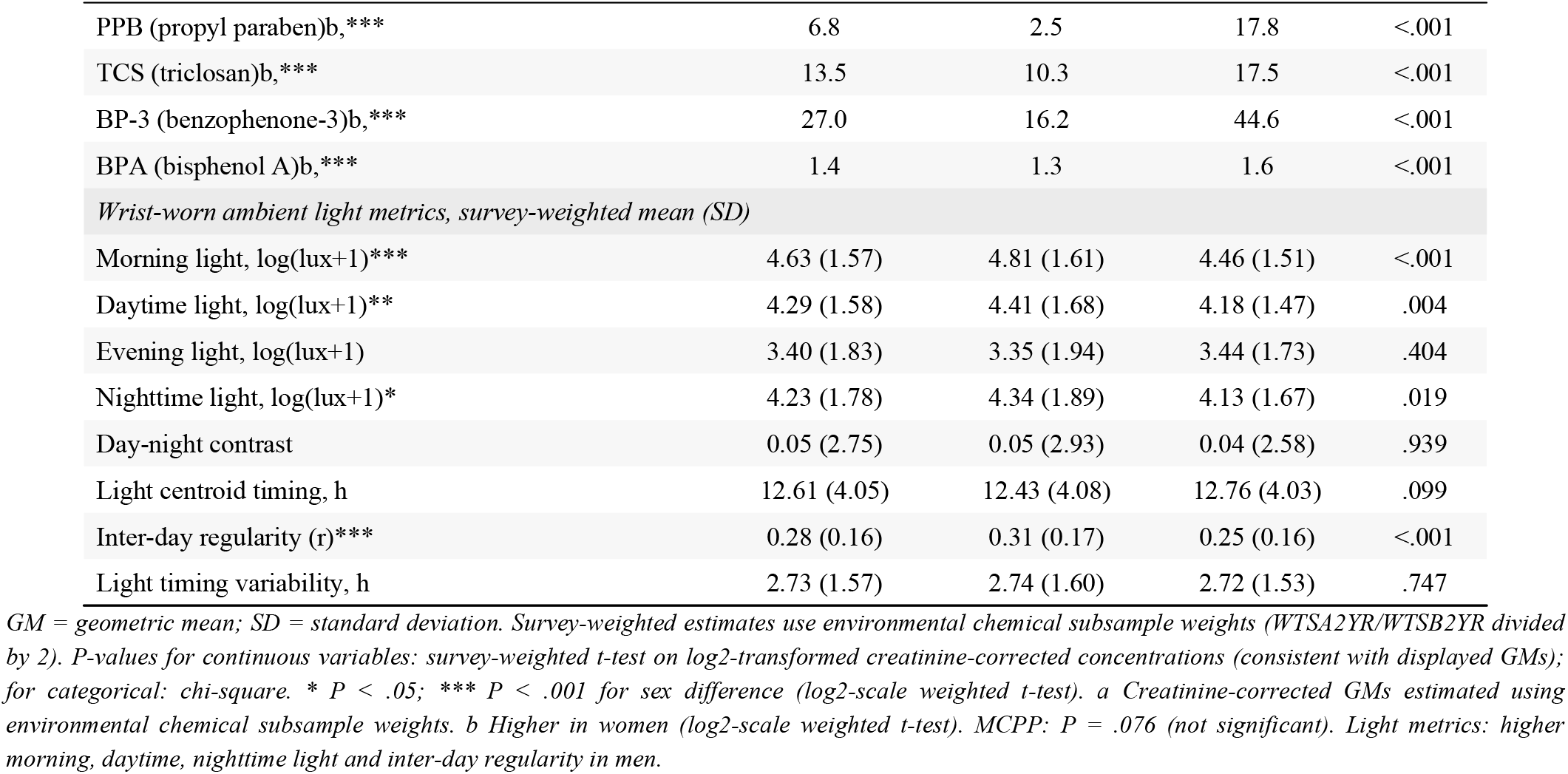
Characteristics of the analytic sample, NHANES 2011-2014 (N = 1,666).

### 3.2 Urinary Biomarker Mixture Index and Wrist-Worn Light Outcomes

Higher urinary biomarker mixture index was associated with three wrist-worn light outcomes after FDR correction (Table 2, Figure 1): greater morning light (beta = +0.54; 95% CI: 0.14, 0.94; FDR q = .028), greater nighttime light (beta = +0.55; 95% CI: 0.21, 0.89; FDR q = .015), and earlier light centroid timing (beta = −1.37 h; 95% CI: −2.14, −0.59; FDR q = .014). Daytime light, day-night contrast, inter-day regularity, and light timing variability showed no significant associations. Evening light was FDR-significant in regression (FDR q = .040) but not directionally consistent in the sensitivity analysis and is treated as exploratory.

**Table 2.** Survey-weighted regression associations between urinary biomarker mixture index and wrist-worn light outcomes, NHANES 2011-2014 (N = 1,666).

| Outcome | beta | 95% CI | P | FDR q | Interpretation |
| --- | --- | --- | --- | --- | --- |
| **Morning light, log(lux+1)** | **+0.54** | **0.14, 0.94** | **.011** | **.028** | **FDR-significant** |
| Daytime light, log(lux+1) | +0.005 | -0.32, 0.33 | .975 | .975 | Not significant |
| Evening light, log(lux+1) | -0.40 | -0.72, -0.07 | .020 | .040 | Exploratory |
| **Nighttime light, log(lux+1)** | **+0.55** | **0.21, 0.89** | **.004** | **.015** | **FDR-significant** |
| Day-night contrast | -0.51 | -0.99, -0.03 | .038 | .060 | Nominal only |
| **Light centroid timing, h** | **-1.37** | **-2.14, -0.59** | **.002** | **.014** | **FDR-significant** |
| Inter-day regularity | +0.010 | -0.011, 0.031 | .340 | .454 | Not significant |
| Light timing variability, h | -0.049 | -0.28, 0.18 | .677 | .774 | Not significant |

beta = coefficient per 1-SD increase in mixture index. Models adjusted for age, sex, race/ethnicity, PIR, education, BMI, log2-cotinine, sleep duration, season. Survey weights: WTS2YR/WTSB2YR / 2. FDR = Benjamini-Hochberg across 8 outcomes.

**Figure 1.**
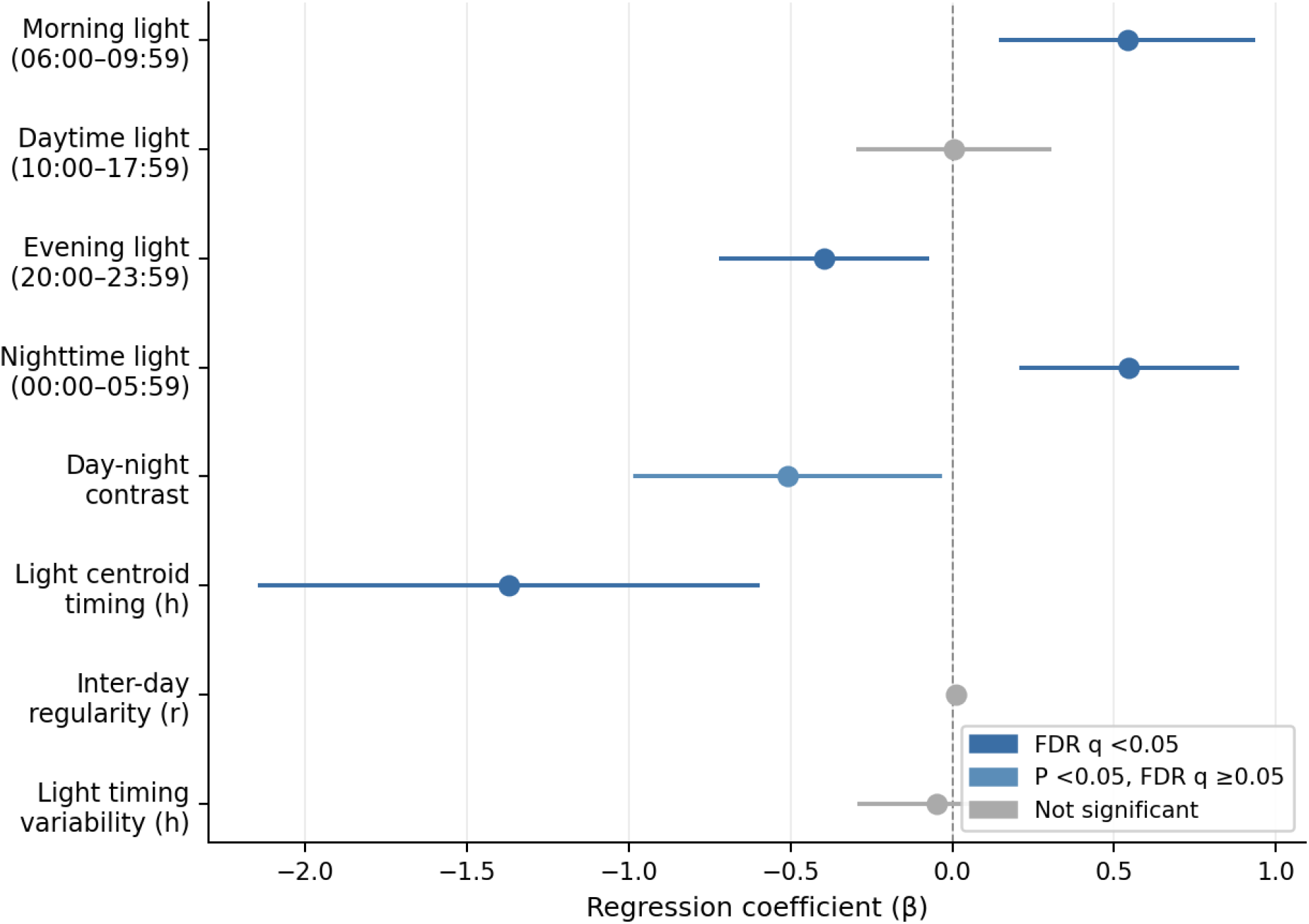
Survey-weighted regression associations (beta, 95% CI) between urinary biomarker mixture index and eight wrist-worn ambient light outcomes, NHANES 2011-2014 (N = 1,666). Dark blue = FDR q < .05; steel blue = nominal P < .05; gray = not significant. Dashed line = null.

### 3.3 Weighted Sensitivity Analysis: Quantile G-Computation

In a weighted sensitivity analysis using quantile g-computation, associations were directionally consistent for the three primary outcomes (Table 3, Figure 2). A joint one-quartile increase across all 15 components was associated with greater morning light (psi = +0.51; 95% CI: 0.35, 0.67; FDR q < .001), greater nighttime light (psi = +0.50; 95% CI: 0.31, 0.69; FDR q < .001), and earlier light centroid timing (psi = −1.05 h; 95% CI: −1.46, −0.63; FDR q < .001). These results use case weights only and should be interpreted as directional corroboration. For light centroid timing, MBP had the largest positive model weight (0.40) and CNP the second (0.37); MCPP had the largest negative weight (0.25).

**Table 3.** Quantile g-computation joint mixture associations with wrist-worn light outcomes, NHANES 2011-2014 (N = 1,666).

| Outcome | psi | 95% CI | P | FDR q | Interpretation |
| --- | --- | --- | --- | --- | --- |
| **Morning light, log(lux+1)** | **+0.51** | **0.35, 0.67** | **<.001** | **<.001** | **Consistent** |
| Daytime light, log(lux+1) | +0.11 | -0.05, 0.27 | .168 | .257 | Not significant |
| Evening light, log(lux+1) | -0.12 | -0.35, 0.11 | .299 | .299 | Not significant |
| **Nighttime light, log(lux+1)** | **+0.50** | **0.31, 0.69** | **<.001** | **<.001** | **Consistent** |
| Day-night contrast | -0.35 | -0.64, -0.06 | .018 | .035 | Sensitivity only |
| **Light centroid timing, h** | **-1.05** | **-1.46, -0.63** | **<.001** | **<.001** | **Consistent** |
| Inter-day regularity | +0.011 | -0.006, 0.028 | .193 | .257 | Not significant |
| Light timing variability, h | -0.096 | -0.26, 0.07 | .258 | .295 | Not significant |

$\psi$ = joint mixture association per one-quartile increase across all 15 components (qqcomp.noboot, $q = 4$ ). Case weights only (WTS2YR/WTSB2YR / 2); variance does not account for complex survey design. FDR = Benjamini-Hochberg.

**Figure 2.**
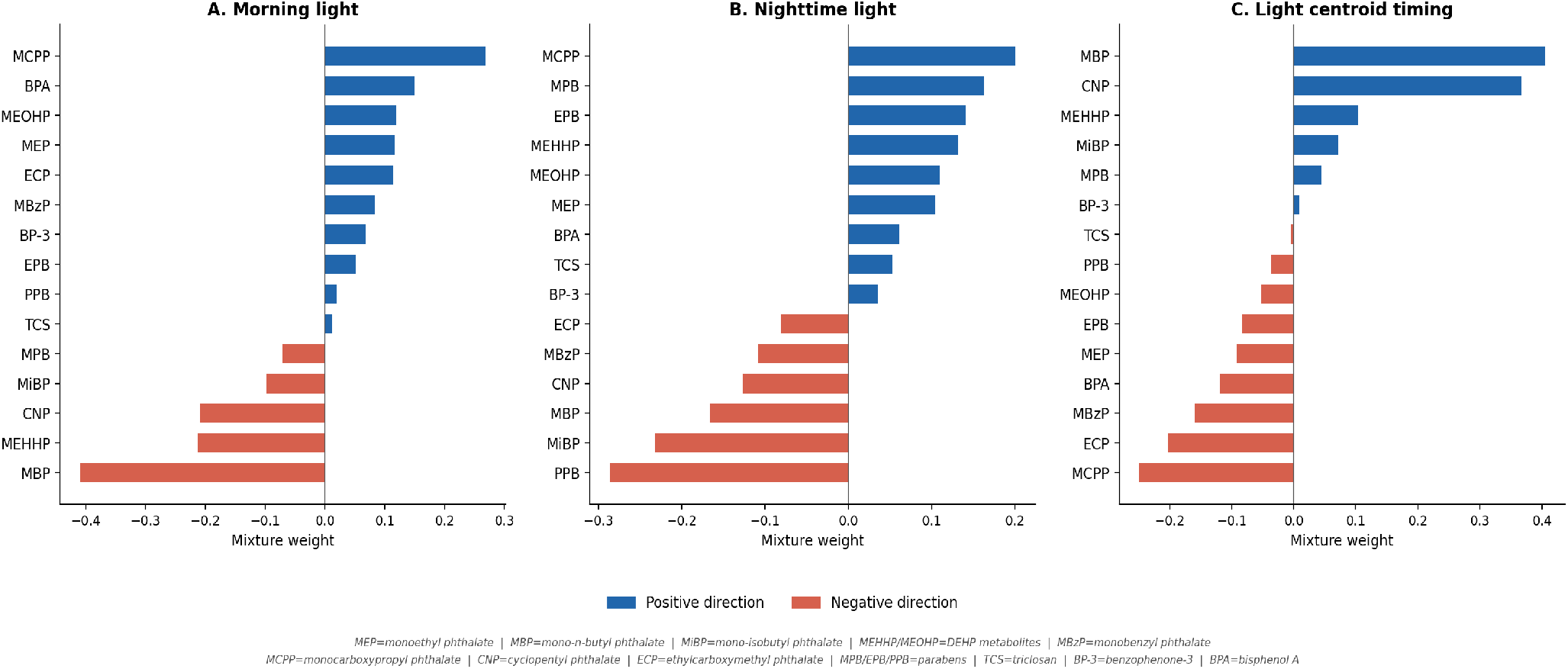
Quantile g-computation component model weights for three outcomes with directionally consistent associations, NHANES 2011-2014 (N = 1,666). Blue = positive weight; red = negative weight. Weights reflect contributions within the fitted model; not individual effect estimates.

### 3.4 Sex-Stratified Analyses

No significant sex modification was observed for any primary outcome (all interaction P > .23; Figure 3). The light centroid association was similar in men (beta = −1.31 h; 95% CI: −2.20, −0.42) and women (beta = −1.40 h; 95% CI: −2.32, −0.47), despite substantially higher creatinine-corrected paraben concentrations in women (PPB GM: males 2.5, females 17.8 ug/g creatinine), suggesting the co-patterning is not driven primarily by sex-differentiated personal care product use.

**Figure 3.**
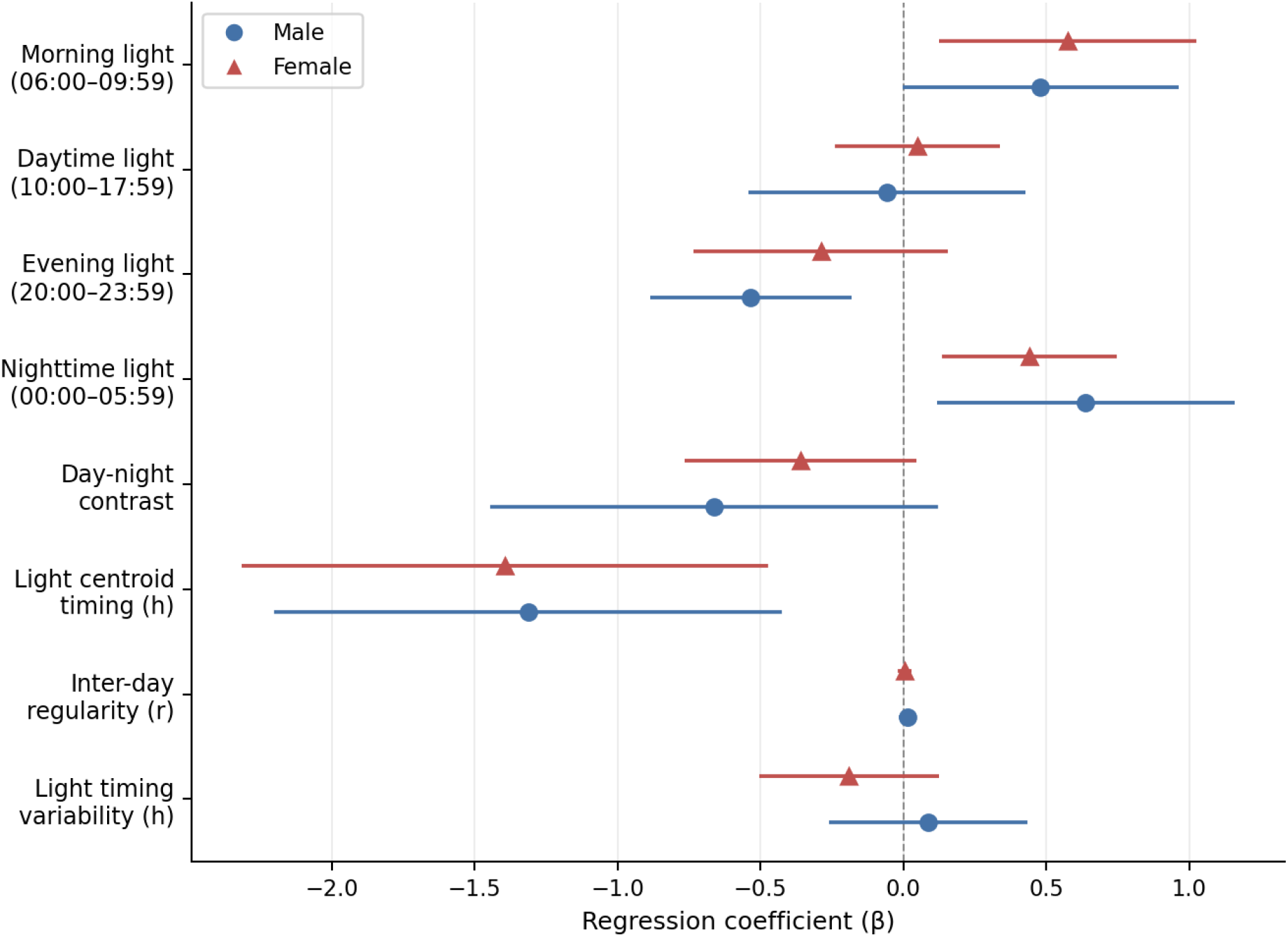
Sex-stratified regression coefficients for the association between urinary biomarker mixture index and eight wrist-worn light outcomes, NHANES 2011-2014. Blue circles = men; red triangles = women. No significant sex interaction (all P > .23).

### 3.5 Chemical Correlation Structure

Pairwise Pearson correlations among the 15 urinary biomarkers (log2-creatinine-corrected) are shown in Figure 4. Parabens (MPB, EPB, PPB) formed a strongly correlated subcluster (r = 0.72-0.89), as did DEHP metabolites MEHHP and MEOHP (r = 0.92). Phthalate-paraben cross-correlations were generally weak to moderate (r = 0.10-0.35), indicating that the 15 biomarkers do not represent a single unified exposure dimension.

**Figure 4.**
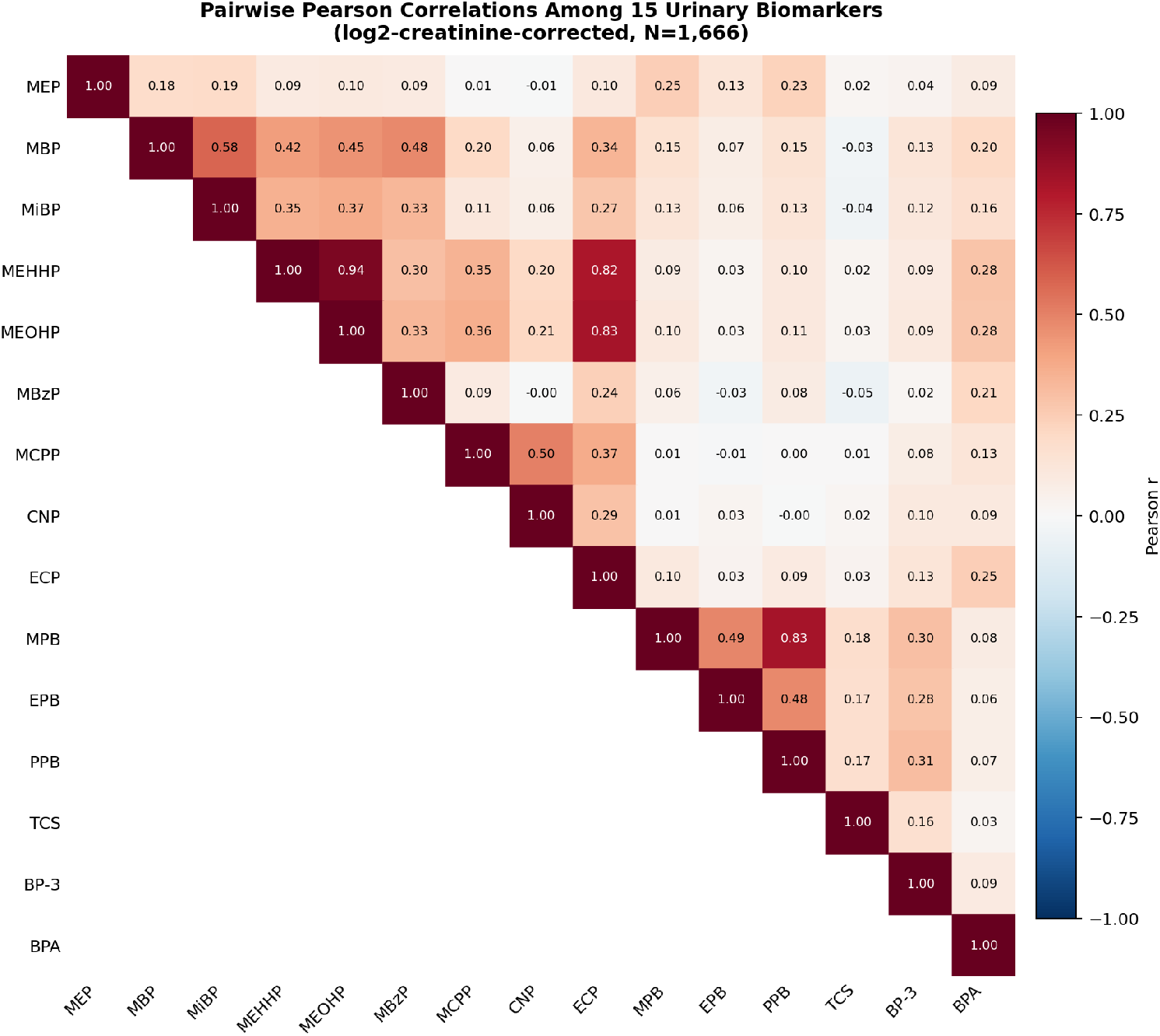
Heatmap of pairwise Pearson correlations among 15 urinary biomarkers (log2-creatinine-corrected), NHANES 2011-2014 (N = 1,666). Blue = positive; red = negative.

## 4 Discussion

In this cross-sectional analysis of 1,666 U.S. adults from NHANES 2011-2014, higher urinary biomarker mixture index was associated with more morning light, more nighttime light, and earlier light centroid timing after FDR correction, with directionally consistent findings in a weighted sensitivity analysis. Associations were consistent across sexes. These findings should be interpreted as short-term co-patterning between urinary consumer-product chemical biomarkers and wrist-worn ambient light exposure, not as evidence that chemical exposures alter circadian light behavior.

The pattern describes a relatively early-timed wrist-level light exposure profile. More morning light and elevated nighttime light are consistent with early rising and early daily activity schedules. Earlier light centroid timing indicates that the lux-weighted center of gravity of light exposure occurs earlier in the day among those with higher mixture burden.

The most plausible explanation for these associations is shared occupational and behavioral pathways. Shift work and early-schedule occupations may simultaneously produce earlier light exposure schedules and higher urinary chemical concentrations through workplace chemical contact, use of personal care products before early shifts, and dietary patterns associated with early schedules [12,13]. The absence of adjustment for employment status, occupation, or shift work is accordingly the most important limitation of this study. The observed associations may be substantially or entirely explained by unmeasured occupational and work-schedule factors.

The quantile g-computation model weight decomposition identified MBP and CNP as having the largest positive model weights for light centroid timing. MBP and CNP are found in industrial solvents, PVC flooring, and adhesives [10], suggesting occupational or environmental sources beyond personal care products. These model weights reflect relative contributions within the mixture model parameterization and should not be interpreted as chemical-specific causal effects.

### 4.1 Limitations

The cross-sectional design precludes causal inference [14]. A single spot urine sample reflects recent rather than chronic exposure due to the short half-lives (hours to days) of phthalates, parabens, phenols, triclosan, and BP-3 [8,9].

Wrist-worn lux represents wrist-level ambient light, not retinal illuminance or melanopic circadian stimulus. Lux does not capture spectral composition relevant to circadian photoreception [7]. The mean nighttime log(lux+1) of 4.23 (approximately 68 lux) likely reflects wakefulness during sleep in addition to environmental nighttime light.

Light centroid timing uses arithmetic averaging of clock hours, which does not account for the circular nature of time. Participants with light exposure near midnight could have distorted estimates, though sensitivity analyses indicated fewer than 2% of participants were affected.

The most critical limitation is the absence of adjustment for employment status, occupation category, and shift work schedule. These factors are the most plausible shared determinants of both urinary chemical profiles and light exposure patterns under this study’s hypothesized mechanism. The observed associations may be substantially or entirely explained by unmeasured occupational confounding.

The urinary biomarker mixture index uses equal weighting across 15 chemically heterogeneous compounds. The analytic sample represents approximately 15% of eligible NHANES adults, reflecting intersection of the PAM and environmental chemical subsamples; survey weighting may not fully address differential selection.

## 5 Conclusions

Higher urinary biomarker mixture co-occurred with earlier-timed wrist-worn ambient light exposure patterns in a nationally representative U.S. adult sample. These associations reflect short-term co-patterning and are most plausibly attributable to shared occupational and behavioral determinants. They should not be interpreted as evidence of a biological effect of chemical exposure on circadian behavior. Future studies with repeated biomarker measurements, objective occupational and shift work data, validated circadian light metrics, and longitudinal designs are needed.

## Author Contributions

A.W.: conceptualization, formal analysis, methodology, software, visualization, writing (original draft). L.Y.: investigation, validation, writing (review and editing). C.L.: investigation, writing (review and editing). A.P.: investigation, writing (review and editing). Y.C.: investigation, writing (review and editing). All authors approved the final manuscript.

## Data Availability

NHANES 2011-2014 data are publicly available at https://www.cdc.gov/nchs/nhanes/. Analytic code is available upon reasonable request to the corresponding author.

## Acknowledgments

The authors thank NHANES participants and staff. NHANES is conducted by the National Center for Health Statistics, CDC. The findings are those of the authors and do not represent the official position of the NCHS or CDC.

